# Plasma extracellular vesicle synaptic proteins as biomarkers of clinical progression in patients with Parkinson’s disease: A follow-up study

**DOI:** 10.1101/2023.05.27.23290639

**Authors:** Chien-Tai Hong, Chen-Chih Chung, Ruan-Ching Yu, Lung Chan

## Abstract

Synaptic dysfunction plays a key role in Parkinson’s disease (PD), and plasma extracellular vesicle (EV) synaptic proteins are emerging as biomarkers for neurodegenerative diseases. This study assessed the efficacy of plasma EV synaptic proteins as biomarkers in PD and their association with disease progression. In total, 144 participants were enrolled, including 101 people with PD (PwP) and 43 healthy controls (HCs). The changes in plasma EV synaptic protein levels between baseline and 1-year follow-up did not differ significantly in both PwP and HCs. In PwP, the changes in plasma EV synaptic protein levels were significantly associated with the changes in unified PD rating scale (UPDRS) part II and III scores. Moreover, PwP with elevated levels (first quartile) of any one plasma EV synaptic proteins (synaptosome-associated protein 25, growth-associated protein 43 or synaptotagmin-1) had significantly greater disease progression in UPDRS part II score and the postural instability and gait disturbance subscore in UPDRS part III than did the other PwP after adjustment for age, sex, and disease duration. These results indicate the promising potential of plasma EV synaptic proteins as clinical biomarkers of disease progression in PD. However, a longer follow-up period is warranted to confirm their role as prognostic biomarkers.

## Introduction

Parkinson’s disease (PD) is the second most common neurodegenerative disease [1] that is well known for its progression, which involves increased disability and burden [2]. Worsening is noted not only in motor symptoms but also in nonmotor ones, particularly cognition. The rate of disease progression varies among people with PD (PwP). In the ongoing Parkinson’s Progression Markers Initiative cohort study, approximately one-third of untreated PwP exhibited rapid progression during the first 2 years of follow-up. By contrast, the remaining PwP had a slow progressive course [3]. Unfortunately, no disease modifying therapy for halting disease progression and no predictor for assessing disease progression are available.

Synapses are sites of neuronal communication, and synaptic degeneration is an early functional pathogenic event in neurodegenerative diseases such as Alzheimer’s disease (AD) and PD [4, 5]. Postmortem studies have revealed a substantial loss of dopamine terminals in the putamen and amygdala [6, 7]. In addition, the loss of glutamatergic corticostriatal synapses has been reported [8]. Mitochondrial dysfunction–related metabolic burden accounts for part of the synaptic loss in PD, and the aggregation of α-synuclein, a synaptic protein that is the major pathognomonic protein in PD, contributes to the pathogenesis of PD [9]. Assessment of synaptic proteins in the cerebrospinal fluid (CSF) can reflect synaptic loss in patients with neurological diseases and is a key area of research interest. Substantial research efforts have been focused on assessing the CSF synaptic pathology to improve the diagnosis of neurodegenerative diseases at an early stage, before neuronal loss, and to monitor clinical progression [10-12]. Several synaptic proteins are promising biomarkers of synaptic function. Synaptosome-associated protein 25 (SNAP-25) is a presynaptic protein that plays a crucial role in neuronal survival, vesicular exocytosis, and neurite outgrowth [13]. Increased CSF levels of SNAP-25 have been reported in PwP [12]. In addition, growth-associated protein 43 (GAP-43) is a presynaptic protein anchored to the cytoplasmic side of the presynaptic plasma membrane [14]. CSF levels of GAP-43 have been reported to be significantly higher in patients with AD than in HCs [15]. Synaptotagmin-1 is a calcium sensor vesicle protein that is vital for rapid synchronous neurotransmitter release in hippocampal neurons [16].

Significantly increased CSF levels of synaptotagmin-1 have been reported in patients with AD and mild cognitive impairment [17]. Regarding PD, one study reported an increase in the level of CSF SNAP-25 but not Ras-related protein 3A or neurogranin. Moreover, treated PwP exhibited higher CSF levels of SNAP-25 than did their drug-naïve counterparts [18].

However, the CSF collection process is moderately invasive, is inevitable during clinical assessment, and can result in some side effects, such as postpuncture headaches. Although studies have assessed blood biomarkers for PD diagnosis and progression, the results are conflicting. A lack of correlation between the peripheral blood content and the brain because of the blood–brain barrier (BBB) is a major obstacle to the identification of blood biomarkers for neurodegenerative diseases [19]. Assessment of peripheral blood extracellular vesicle (EV) proteins can be an alternative approach. EVs are tiny vesicles covered with a lipid membrane. They contain proteins, lipids, and nucleic acid responsible for cell-to-cell signal transmission. The integrity of EVs can be maintained when crossing the BBB [20]. Plasma EV biomarkers being rapidly developed for PD [21]. EV-cargo α-synuclein has been the most studied target that has exhibited strong potential for distinguishing PwP from HCs and other patients with atypical parkinsonism [22-26]. EV-cargo tau, β-amyloid, neurofilament light chain, brain-derived neurotrophic factor, and insulin receptor substrate have also been assessed in PwP [27-31]. These results indicate the potential role of EV content as biomarkers in PD.

The levels of synaptic proteins in blood exosomes, a specific type of EV, decrease in patients with AD and frontotemporal dementia [32]. Moreover, blood exosomal SNAP-25, GAP-43, neurogranin, and synaptotagmin-1 levels are lower in patients with AD. A combination of exosomal synaptic protein biomarkers could predict cognitive impairment [33]. Regarding PD, a cross-sectional study also had demonstrated that the synaptic proteins inside the blood neuron-derived exosomes are reduced in PwP compared with healthy controls (HCs), which can distinguish PwP with HCs with around 80% accuracy[34]. Considering the plasma EVs remain stable up to 90 days [35]; this prevents the fluctuation of free-form synaptic proteins because of transient surge or degradation. Moreover, SNAP-25 is transported in the blood by EVs [36]. However, there is no information from the cohort study of PD. Therefore, this study assessed the association between plasma EV synaptic proteins and PD progression and determined whether plasma EV synaptic proteins could be used as clinical biomarkers to predict the progression of PD.

## Results

The participants’ demographic data at baseline and 1-year follow-up are presented in Table 1. In total, 144 participants (101 PwP and 43 HCs) were followed up. No significant difference was noted in plasma EV SNAP-25, GAP-43, and synaptotagmin-1 levels at baseline and follow-up between PwP and HCs after adjustment for age and sex (Figures 1A [representative image] and B–D [dot plot]) (Supplementary Figure 1 for the original blot of the representative image).

**Table 1.** Demographic data of study participants.

|  | HCs, n = 43 | PwP, n = 101 |
| --- | --- | --- |
| Age (years) | 65 (10.24) | 69 (7.76) |
| Women | 15 | 48 |
| Baseline |  |  |
| MMSE | 27 (3.92) | 26 (4.15) |
| MoCA | 23 (4.63) | 21 (5.70) |
| Disease duration (years) | - | 2 (2.24) |
| UPDRS-II | - | 8 (5.58) |
| UPDRS-III | - | 22 (9.30) |
| 1-year follow-up |  |  |
| MMSE | 28 (4.12) | 27 (5.61) |
| MoCA | 24 (5.82) | 23 (6.45) |
| UPDRS-II | - | 11 (6.41) |
| UPDRS-III | - | 19 (9.39) |

Data was presented as median (standard deviation). HC, healthy control; PwP, people with Parkinson's disease; MMSE, Mini-Mental State Examination; MoCA, Montreal cognitive assessment; UPDRS, unified Parkinson's disease rating scale.

**Figure 1.**
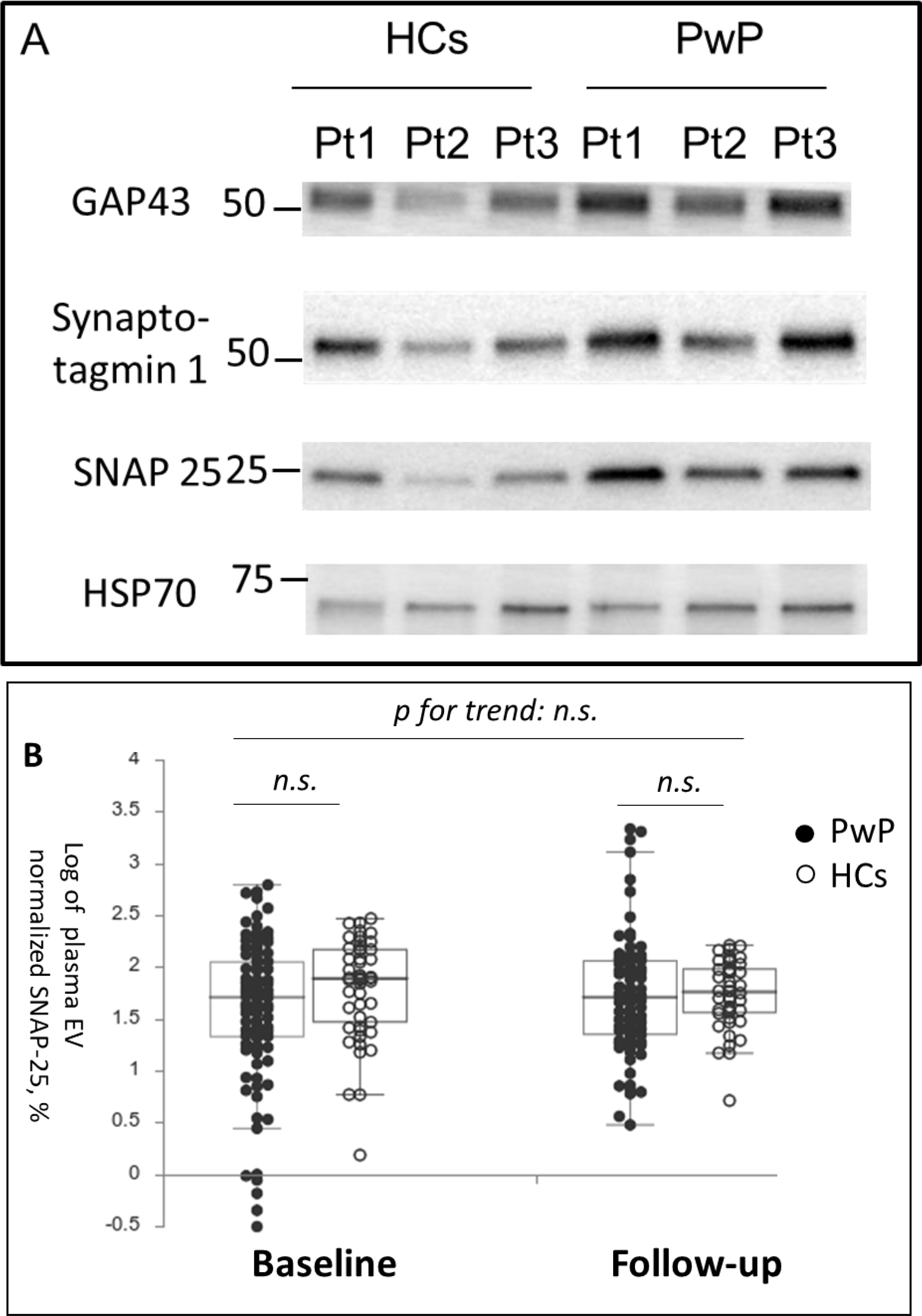
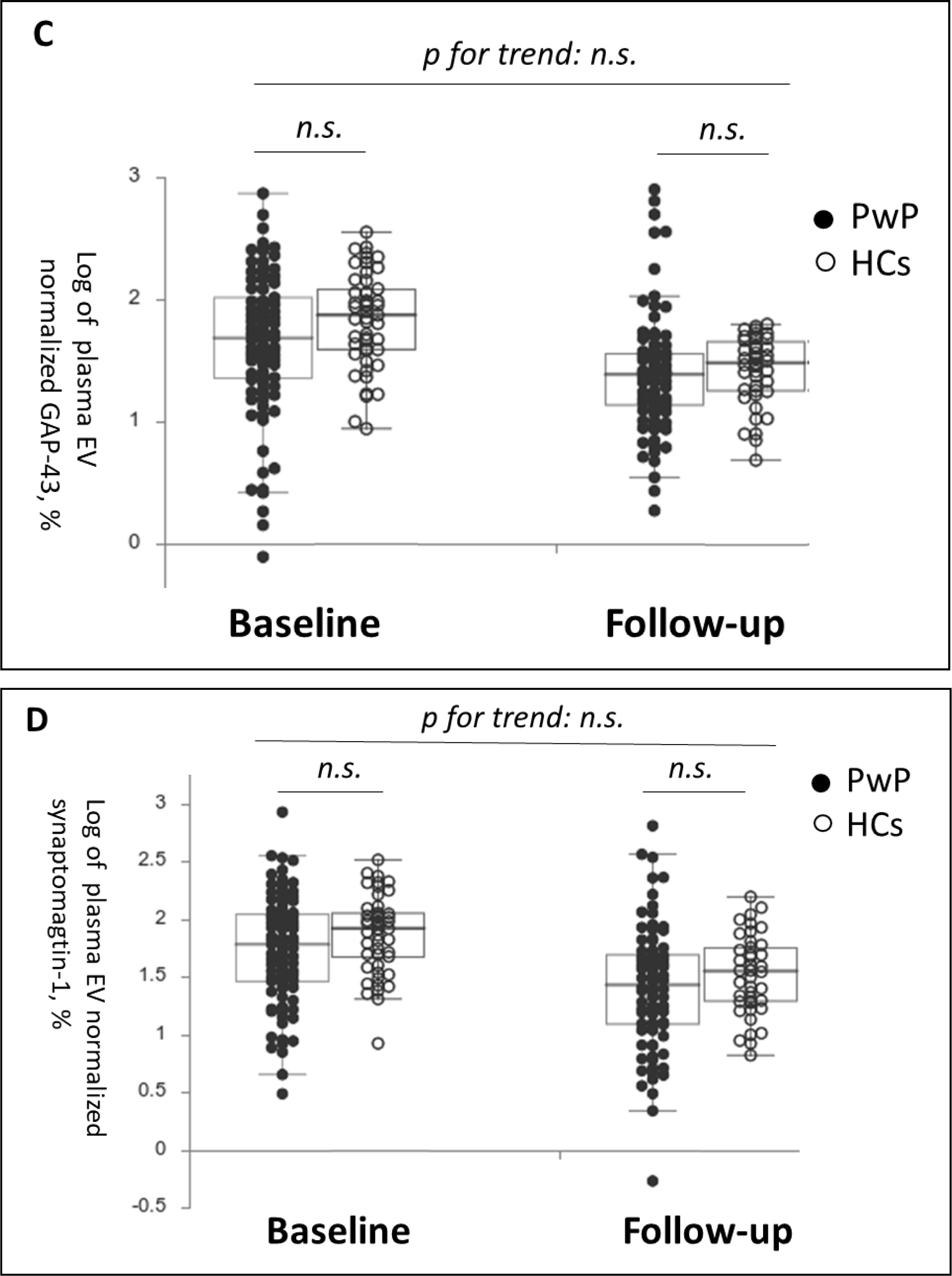
Baseline and follow-up synaptic protein levels in plasma extracellular vesicles (EVs) between patients with Parkinson’s disease (PwP) and healthy controls (HCs). (A) Representative protein blot images of different synaptic proteins, including SNAP-25, GAP-43, and synaptotagmin-1. Heat shock protein 70 (HSP-70) was the protein loading control. (B–D) Comparison of plasma SNAP-25, GAP-43, and synaptotagmin-1 levels between PwP and HCs at baseline and follow-up. Data are presented using a dot plot displaying the median and first and third quartile values. n.s., non-significant.

We assessed the association between the changes in plasma EV synaptic proteins levels and the changes in clinical parameters in PwP through the generalized linear model (Table 2). The changes in the total score of unified Parkinson disease rating scale (UPDRS)-II was positively associated with the change of plasma EV synaptic proteins (SNAP-25, GAP-43, and synaptotagmin-1); the change of total score of UPDRS-III and akinetic rigidity (AR) subscore were significantly associated with the change of plasma EV GAP-43 and synaptotagmin-1. The changes in mini-mental status examination (MMSE) and Montral cognitive assessment (MoCA) scores were non-significantly associated with the changes in plasma EV synaptic protein levels.

**Table 2.**
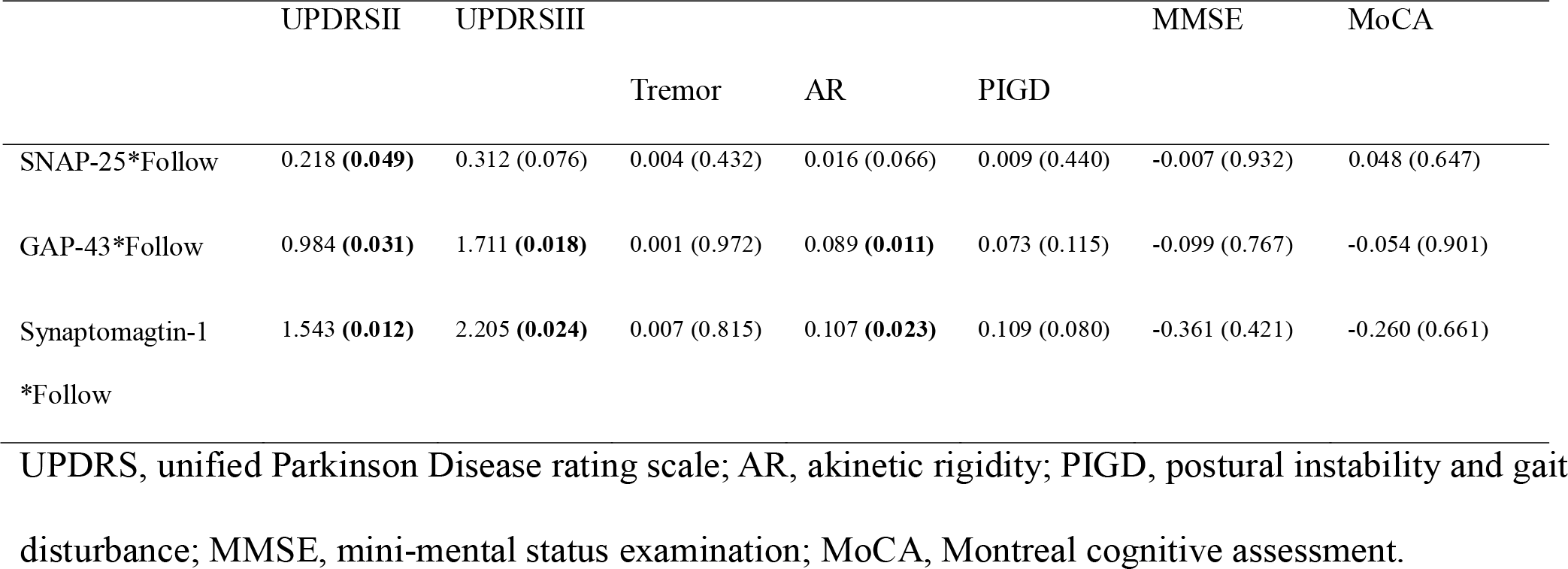
The association between the change of plasma EV synaptic proteins abundance (between baseline and follow-up) with the change of clinical severity in motor and cognitive domains (between baseline and follow-up) in people with Parkinson’s disease. A generalized linear model was employed and the data was presented as coefficient (p value).

We further assessed the association between the severity of the clinical parameters of PD at follow-up and the baseline plasma EV SNAP-25, GAP-43, and synaptotagmin-1 levels. After adjustment for age, sex, and disease duration, the plasma EV SNAP-25, GAP-43, and synaptotagmin-1 levels were nonsignificantly associated with the UPDRS-II, UPDRS-III, MMSE, and MoCA scores at follow-up (Figure 2; for details, refer to Supplementary Table 1). However, after we categorized the UPDRS-III scores into tremor, AR, and postural instability and gait disturbance (PIGD) subscores, the baseline plasma EV SNAP-25 and GAP-43 levels exhibited a significant positive correlation with the PIGD subscores at follow-up; a similar trend was noted in the plasma EV synaptotagmin-1 level.

**Figure 2.**
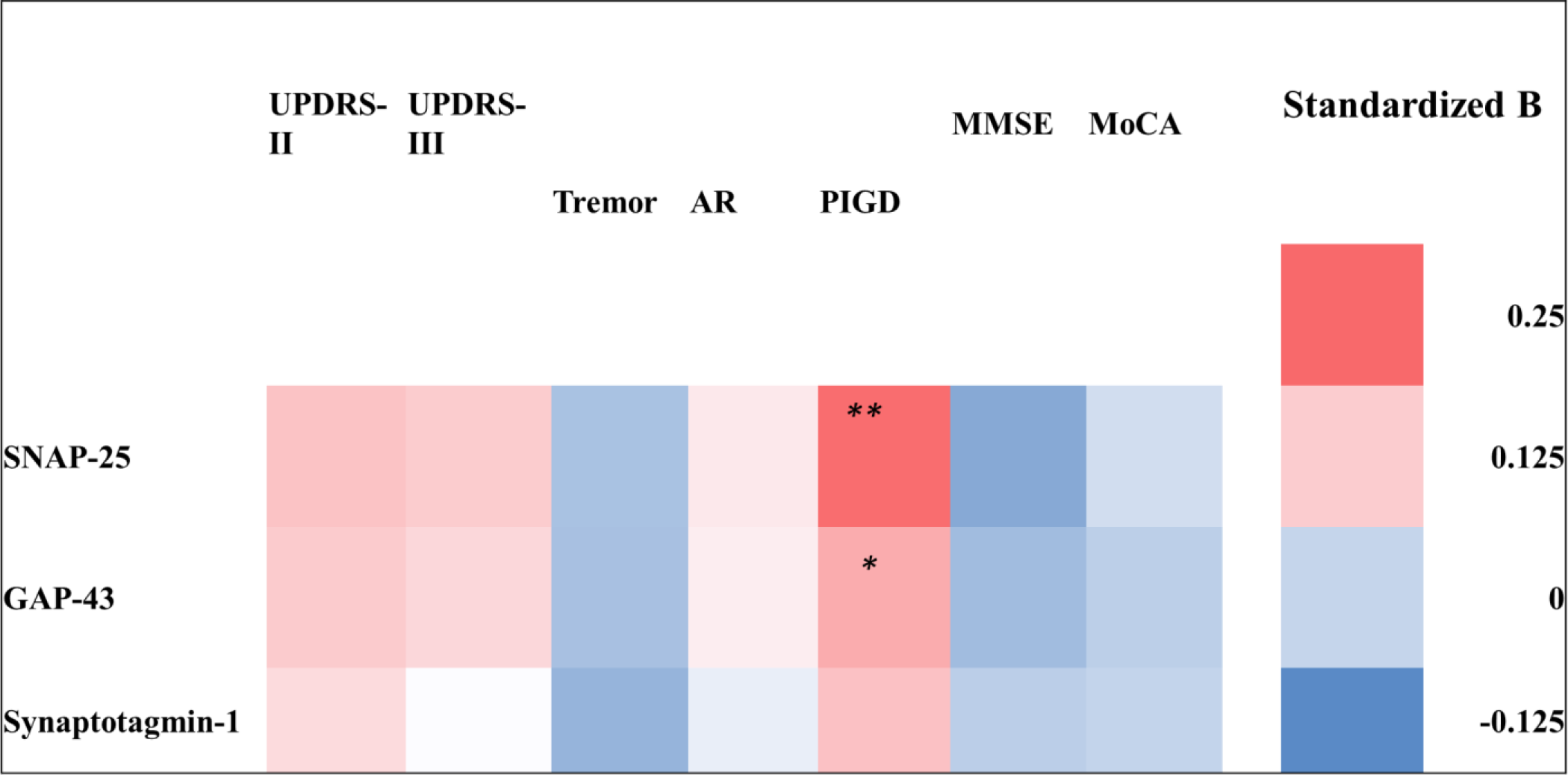
Heatmap of the association between baseline plasma extracellular vesicle (EV) synaptic protein levels and clinical assessment parameters at follow-up in patients with Parkinson’s disease (PwP). The logistic regression model was used to assess the baseline plasma EV SNAP-25, GAP-43, and synaptotagmin-1 levels. The motor symptoms were assessed based on the Unified Parkinson Disease Rating Scale (UPDRS)-II and UPDRS-III scores and tremor, akinetic rigidity (AR), and postural instability and gait disturbance (PIGD) subscores, and cognitive function was assessed using the Mini-Mental State Examination (MMSE) and Montreal Cognitive Assessment (MoCA) scores. The association is presented using standardized β values. Detailed results of the regression model are provided in Supplementary Table 1. *, p < 0.05; **, p < 0.01.

Finally, we grouped the PwP on the basis of their baseline plasma EV synaptic protein level, with a cutoff at the first quartile. Overall, PwP with elevated baseline levels of plasma EV synaptic proteins had poor total scores of UPDRS-II and UPDRS-III and poor PIGD subscores of UPDRS-III (Table 3 and detail in supplementary Table 2). In contrast, a significant greater improvement in the tremor score was noted in the PwP with elevated baseline levels of plasma EV synaptic proteins. Moreover, PwP with elevated baseline levels of plasma EV synaptic proteins exhibited significantly greater deterioration, as assessed using the UPDRS-II scores and PIGD subscores in UPDRS-III. After adjustment for age, sex, and disease duration, repeated-measures analysis of covariance revealed that the estimated marginal means of UPDRS-II scores and PIGD subscores of PwP with elevated levels of any one plasma EV synaptic protein were significantly worse at follow-up (but not at baseline) than those of PwP without elevated plasma EV synaptic protein levels, indicating significantly faster deterioration in the former patient group (Figure 3).

**Table 3.**
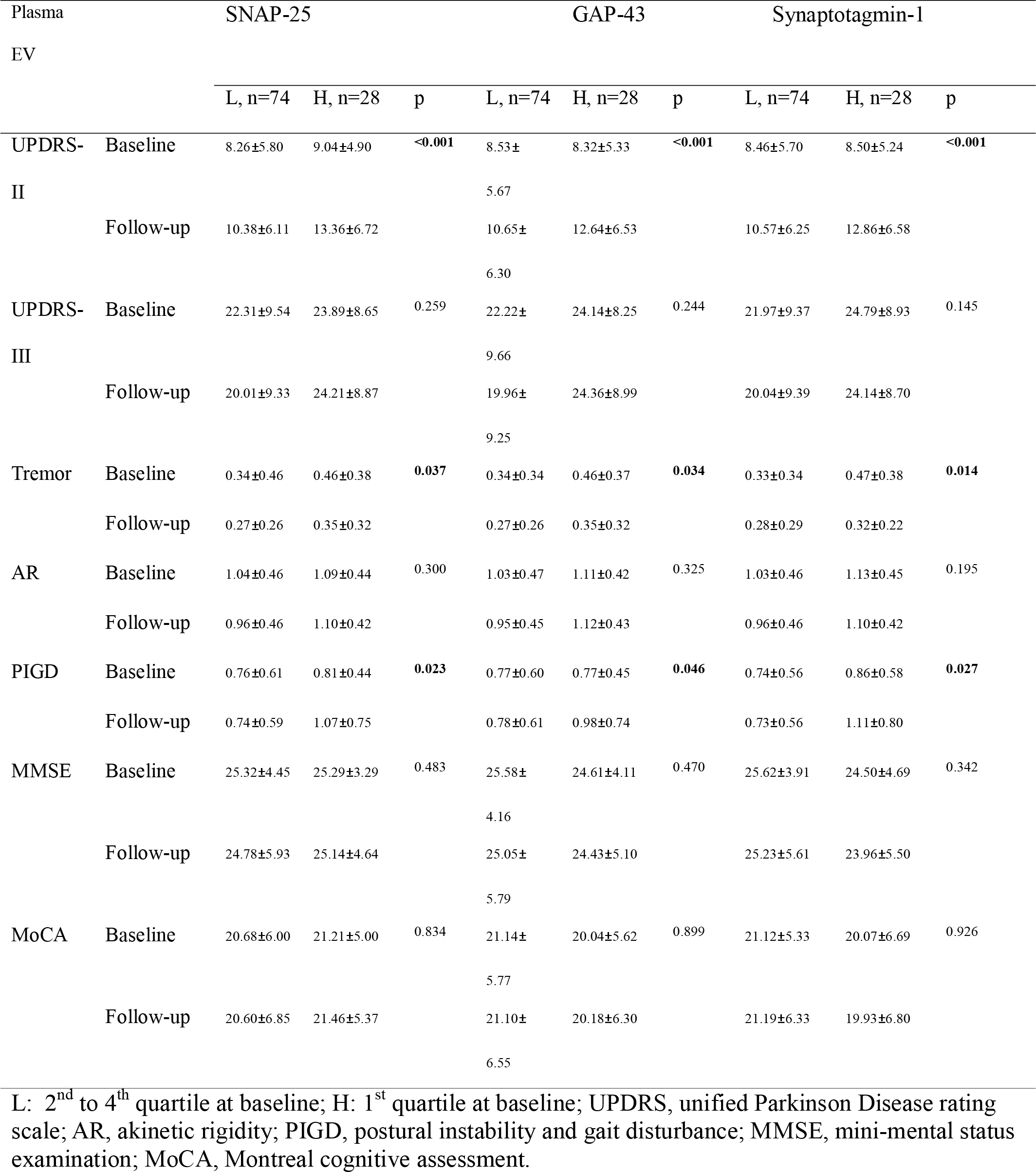
The clinical severity in people with Parkinson’s disease with and without elevated (1^st^ quartile) baseline plasma extracellular vesicle (EV) synaptosome-associated protein 25 (SNAP-25), growth-associated protein 43 (GAP-43) and synaptotagmin-1. P value indicated the inter-group comparisons for the changes.

**Figure 3.**
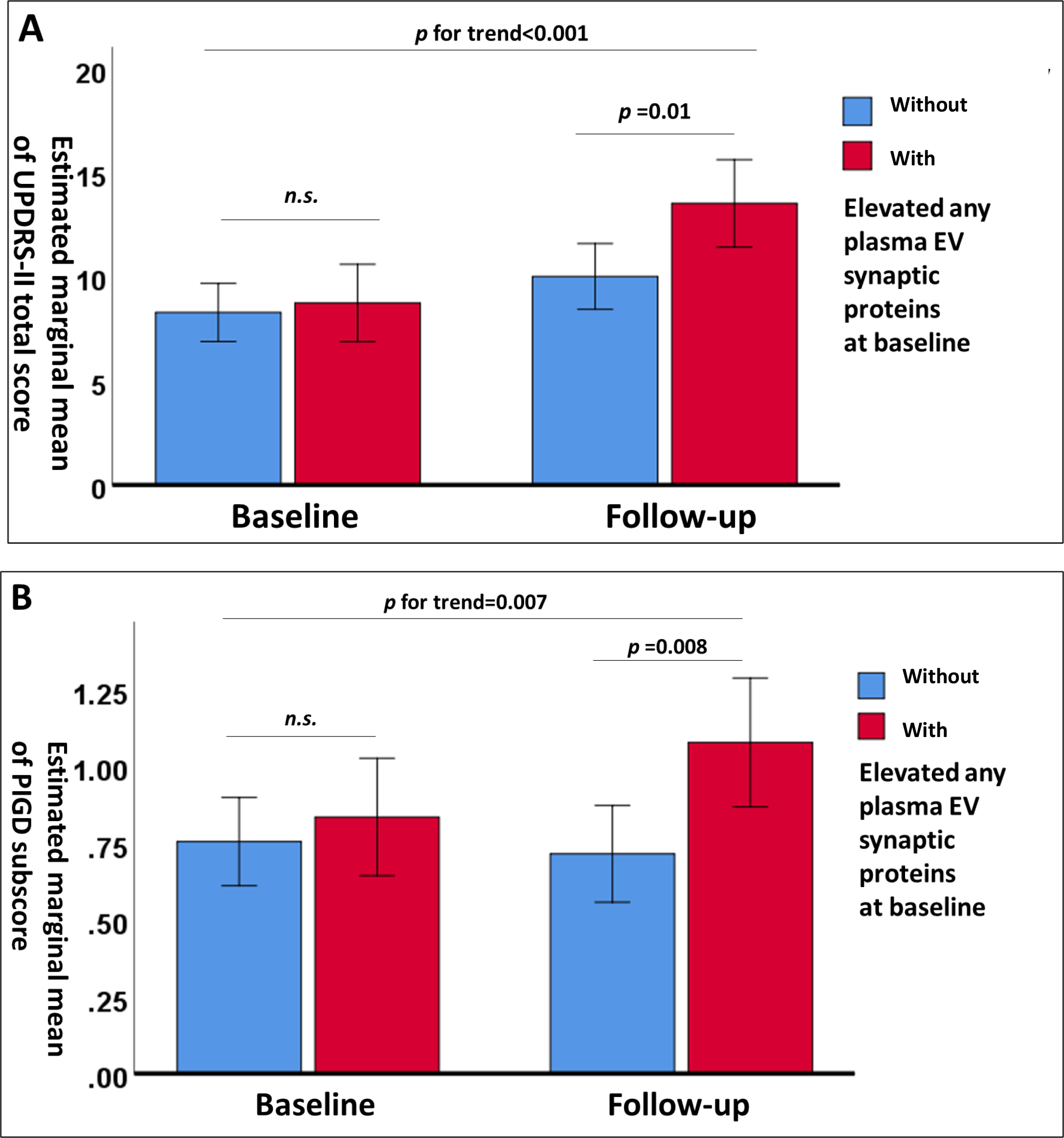
Changes in estimated marginal means of Unified Parkinson Disease Rating Scale (UPDRS)-II total scores (A) and postural instability and gait disturbance (PIGD) subscores (B) after adjustment for age, sex, and disease duration in patients with Parkinson’s disease (PwP) with and without elevated levels of any one plasma extracellular vesicle synaptic protein (first quartile) at baseline and follow-up. Data are presented as means with 95% confidence intervals. n.s., nonsignificant.

## Discussion

In this study, although no significant difference in plasma EV synaptic protein levels was noted between PwP and HCs, changes in plasma EV synaptic protein levels in PwP were associated with motor decline. In addition, baseline plasma EV synaptic protein levels were associated with clinical outcomes, as assessed using PIGD subscores at follow-up. PwP with elevated baseline levels of any one plasma EV synaptic protein (SNAP-25, GAP-43, or synaptotagmin-1) exhibited significant deterioration in activities of daily living (as assessed by UPDRS-II scores and PIGD subscores in UPDRS-III) between baseline and follow-up. These results indicate the promising potential of plasma EV synaptic proteins as biomarkers for PD, particularly its progression.

Significant worsening of UPDRS-II scores and PIGD subscores in UPDRS-III was noted in PwP with higher baseline levels of plasma EV synaptic proteins (first quartile). UPDRS-II is a crucial but underestimated parameter for assessing PwP; it can be used to assess the daily functional capability related to motor symptoms in PwP without temporary drug interruptions, as in the case of UPDRS-III [37, 38]. Assessment of the motor subtypes of PD revealed that the PIGD subtype is associated with an increased Lewy body and α-synuclein burden; rapid decline; and increased risks of cognitive impairment, falls, and mortality in comparison with the tremor-dominant (TD) subtype, which has a relatively benign course [39-42]. PwP may convert from the TD subtype to the PIGD subtype during disease progression [43]. Moreover, PIGD-related motor symptoms respond to dopaminergic medications to a lesser extent than do tremor-, akinesia-, and rigidity-related symptoms [44], which may more directly reflect the progression of the disease. We observed that PwP with elevated levels of any one plasma EV synaptic protein exhibited greater deterioration, as assessed using UPDRS-II scores and PIGD subscores. High EV synaptic protein levels conventionally indicate increased synaptogenesis; this contrasts the synaptic loss pattern noted in PD. Decreased blood exosomal synaptic protein levels have been reported in patients with AD and frontotemporal dementia [32]. However, in PwP who are at the early disease stage, compensatory synaptic sprouting and increased synaptic plasticity are noted in the striatum. This compensation, also known as motor reserve, may temporarily decrease the clinical disease burden. However, the patient would be more vulnerable to deterioration if the compensation is overshadowed by degeneration [45]. For instance, in one study, PwP who engaged in higher premorbid exercises exhibited milder motor symptoms despite having a similar gradient of striatal dopaminergic reduction at baseline; however, they exhibited more rapid deterioration [46]. Because our study mainly included PwP who had the early stage of disease (mean disease duration of less than 3 years) and a mild disease burden (baseline UPDRS-II score = 8.49), the elevated plasma EV synaptic protein levels may indicate the activation of the compensatory process. Such PwP are at an increased risk of rapid deterioration and disease progression and should be candidates for disease-modifying interventions, including pharmacological and nonpharmacological treatments.

The strength of this study is that it is the first study to assess the changes in plasma EV synaptic protein levels and determine the association between the changes in plasma EV synaptic protein levels and cognitive decline in PwP. Considering the well-established role of synaptic degeneration and plasticity in PD, the theoretical background of the use of these synaptic proteins as plasma EV biomarkers for PD is confirmed. Moreover, most synaptic proteins are neuron derived, thereby preventing contamination from nonneuronal tissues. The significant association between the changes in plasma EV synaptic proteins levels and the changes in UPDRS-II, UPDRS-III and AR subscore suggests the efficacy of these proteins in detecting motor decline in PwP, which can serve as a objective parameter for further disease-modification clinical trial. Elevated baseline plasma EV synaptic protein levels can also predict rapid deterioration in PwP, highlighting the importance of motor reserve and synaptic plasticity in the progression of PD for future research. Although any single synaptic protein cannot precisely predict the progression of PD, these protein targets may be considered for use in the biomarker panel and analyzed using an artificial intelligence–assisted artificial neural network, which is widely used to predict the outcomes of several neurological diseases [47-50].

This study has some limitations. Technically, semiquantitative assessment of plasma EV synaptic protein (SNAP-25, GAP-43, and synaptotagmin-1) levels was performed using western blot analysis. The lack of absolute values, i.e. from the results of enzyme-linked immunosorbent assay, limits further clinical application. In addition, because numerous synaptic proteins are involved in the pathogenesis of PD, the three selected proteins may not reflect all features of the synaptic condition. The selection of HSP-70 as the control for the synaptic proteins quantification of plasma EV is not undisputable. Two types of EV proteins are used for this role: membrane proteins and intravesicular proteins. Membrane proteins include CD9, CD63, and CD81; intravesicular proteins include TSG101, annexins, and chaperone proteins, such as HSP-70. Since the expression of HSP-70 is usually steady, we used the HSP-70 as the internal control, but it is a limitation of the present study. Furthermore, this study evaluated the overall plasma EVs rather than specifically focusing on neuron-derived exosomes, potentially introducing a bias towards somatic-origin EVs. Nonetheless, it is worth noting that synaptic proteins primarily originate from neurons. Even when considering neuron-derived exosomes, it’s important to recognize that they are not exclusively derived from the brain, which can lead to contamination from the peripheral nervous system. The final technical issue in the present study was the relatively small size of the isolated EVs. Despite the primary focus on isolating exosomes, which are the smallest type of EVs, it’s important to consider that the presence of small-sized EVs could potentially be attributed to EV fragmentation that occurs during the freezing and thawing processes. Regarding the limitation from the cohort, it is also worth mentioning that the 1-year follow-up period to assess the progression of PD was relatively short and may have been insufficient to detect significant disease progression. On the other hand, the evaluation of motor symptoms occurred in a hospital setting where we did not ask patients to stop taking their anti-PD medications due to safety concerns like the risk of falls. As a result, specific motor symptoms, particularly tremor and AR, which are more sensitive to medication compared to PIGD, may have been effectively managed by the anti-PD medications. This could potentially explain the improvement in tremor observed between the baseline and one-year follow-up, especially among PwP with elevated baseline plasma EV synaptic proteins. Additionally, synaptic dysfunction is a frequently observed phenomenon in several neurological diseases, and it is not exclusive to PD. Consequently, the HC group in our current study may have included individuals with coexisting neurological conditions, potentially explaining the lack of a significant difference between the PD group and the HCs. However, this approach also illuminates the significance of synaptic dysfunction in the advancement of PD. This insight can be invaluable for monitoring disease progression, particularly in the context of clinical trials focused on disease modification. Choosing the first quartile as a cut-off value of plasma EV synaptic proteins is also one of the limitations of the study. While developing new biomarkers, there was no clear cut-off value as reference for the continuous variable, and percentile is considered for predicting the prognosis[51]. Further studies are required to validate this application. Lastly, the results form a mono-centric, small-scale and short-period PD cohort required further validation

In conclusion, this study revealed that changes in the levels of plasma EV synaptic proteins, namely SNAP-25, GAP-43, and synaptotagmin-1, are associated with motor decline in PwP. Elevated baseline plasma EV synaptic protein levels can predict increased deterioration of motor function, particularly PIGD symptoms, in PwP. Our results indicate that plasma EV synaptic proteins have the potential to be used as biomarkers of PD progression and detection. A longer longitudinal follow-up is warranted to clearly assess the prognostic efficacy of plasma EV synaptic proteins in PwP.

## Methods

### Study participants

This study included 101 PwP and 43 HCs. PD was diagnosed in accordance with the criteria used in another study [52]. Patients diagnosed as having early-to-mid-stage PD (Hoehn and Yahr stage I–III) were invited to participate in this study. HCs were excluded if they had comorbidities, such as neurodegenerative, psychiatric, or major systemic diseases (malignant neoplasm or chronic kidney disease). HCs were mainly recruited from neurological outpatient clinics; they had minor chronic health conditions, such as hypertension, diabetes, or hyperlipidemia. This study was approved by the Joint Institutional Review Board of Taipei Medical University (approval no. N201609017 and N201801043).

### Clinical assessments

The participants’ background data were obtained through a personal interview. Their cognitive function was assessed by trained nurses using the Taiwanese versions of the MMSE and MoCA. The severity of PD was assessed using parts I, II, and III of the UPDRS during clinic visits. PwP were assumed to be in their “on” time. Tremor, AR, and PIGD subscores were calculated from the subitems in UPDRS-III as described previously [53], with some modifications.

### Plasma EV isolation and characterization

Venous blood was collected by from PwP and HCs after their outpatients clinic (non-fasting) by 21 gauge needle, and the plasma was isolated through centrifugation at 13,000 × *g* for 20 minutes immediately. Plasma was storage in the −80°C freezer before EV isolation. Plasma EVs were isolated from 1 mL of plasma by exoEasy Maxi Kit (Qiagen, Valencia, CA, USA), a membrane-based affinity binding step to isolate exosomes and other EVs without relying on a particular epitope, in accordance with the manufacturer’s instructions and storaged in the −80°C freezer. The isolated plasma EVs were then eluted and stored. Usually, 400 μL of eluate is obtained per mL of plasma. The isolated plasma EVs were validated according to the International Society of Extracellular Vesicles guidelines, which include1.markers, including the presence of CD63 (ab59479, Abcam, Cambridge, UK), CD9(ab92726, Abcam, Cambridge, UK), tumor susceptibility gene 101 protein (GTX118736, GeneTex, CA, USA) and negative of cytochrome c (ab110325; Abcam, Cambridge, UK) 2. Physical characterization through the nanoparticle tracking analysis, which demonstrated the majority of the size of EV are mainly within 50-100nm 3. The morphology from the electron microscopy analysis. The validation had been described previously [29-31].

### Quantification of plasma EV synaptic proteins

The isolated plasma EVs were directly lysed using protein sample buffer (RIPA Lysis Buffer, Millipore) and analyzed using protein sodium dodecyl sulfate–polyacrylamide gel electrophoresis. Antibodies against SNAP-25 (GeneTex, GTX113839, 1:1000), GAP-43 (GeneTex, GTX114124, 1:5000), and synaptotagmin-1 (GeneTex, GTX127934, 1:1000) were used for the analysis. The antibodies were prepared in Tris-buffered saline containing 0.1% Tween 20 and 5% bovine serum albumin. Secondary antibodies, including antimouse immunoglobulin G (IgG)-conjugated horseradish peroxidase (HRP; 115-035-003) and antirabbit IgG-conjugated HRP (111-035-003), were purchased from Jackson ImmunoResearch. Protein blot intensities were quantified using ImageJ software. The expression levels of plasma EV synaptic proteins were normalized to that of heat shock protein 70 (Proteintech, Cat.10995-1-AP, 1:2000). For each participant, equal volume of EV suspension (5μl) was applied to the protein quantification. To ensure that the data could be compared between different gels, all the data were normalized to the average of the control group in the same gel.

### Statistical analyses

All statistical analyses were performed using SPSS for Windows 10 (version 26; SPSS Inc., Chicago, IL, USA). A linear mixed model was used to assess whether the changes in plasma EV synaptic protein levels differed between PwP and HCs after adjustment for age and sex. A generalized linear model was used to determine the association between the changes in plasma EV synaptic protein levels and the changes in clinical parameters in PwP after adjustment for age, sex, and disease duration. Multivariate logistic regression was performed to assess the association between plasma EV synaptic proteins and clinical parameters at follow-up in PwP after adjustment for age, sex, and disease duration. Repeated-measures analysis of covariance with estimated marginal means was employed to compare the changes in clinical parameters between baseline and follow-up in PwP with elevated baseline levels (first quartile) of any one plasma EV synaptic protein. Finally, p values < 0.05 were considered statistically significant.

## Data Availability

Please contact the corresponding author (CT Hong). The availability of data and materials requires permission from the TMU-JIRB.

## Declarations

### Ethics approval and consent to participate

This study was approved by the Joint Institutional Review Board of Taipei Medical University (TMU-JIRB approval no. N201609017 and N201801043). Written informed consent was obtained from all participants for participation in the study.

### Consent for publication

All authors have read and approved the final version of the manuscript. All authors agree to the present state of authorship and have signed a statement attesting to the authorship.

### Authors’ contributions

Study conception and design: CT Hong, L Chan, and CC Chung. Data acquisition and analysis: CT Hong, L Chan, and CC Chung. Data interpretation: CT Hong and RC Yu. Manuscript writing and revision: CT Hong, CC Chung, L Chan, and RC Yu. Provision of resources and administrative oversight: CT Hong. All authors have read and approved the final manuscript.

## Acknowledgments

None

