## Supplementary information for "Plasma extracellular vesicle synaptic proteins as biomarkers of clinical progression in patients with Parkinson’s disease: A follow-up study"

Supplementary Table 1. Association between the baseline plasma EV synaptic proteins with the clinical severity in people with Parkinson’s disease at follow-up with the adjustment of age, sex, disease duration and the baseline severity of corresponding item, presented as standardized B and p value.

|  | UPDRSII | UPDRSIII |  |  |  | MMSE | MoCA |
| --- | --- | --- | --- | --- | --- | --- | --- |
|  |  |  | Tremor | AR | PIGD |  |  |
| SNAP-25 | 0.137 (0.132) | 0.126 (0.135) | -0.032 (0.753) | 0.090 (0.278) | **0.216 (0.004)** | -0.069 (0.400) | 0.014 (0.811) |
| GAP-43 | 0.127 (0.162) | 0.111 (0.186) | -0.034 (0.737) | 0.084 (0.312) | **0.166(0.030)** | -0.041 (0.617) | -0.010 (0.862) |
| Synaptomagtin-1 | 0.108 (0.223) | 0.064 (0.41) | -0.055 (0.582) | 0.042 (0.609) | 0.139 (0.06) | -0.009 (0.911) | -0.002 (0.967) |

UPDRS, unified Parkinson Disease rating scale; AR, akinetic rigidity; PIGD, postural instability and gait disturbance; MMSE, mini-mental status examination; MoCA, Montreal cognitive assessment.

Supplementary Table 2 The clinical severity in people with Parkinson’s disease with and without elevated (1st quartile) baseline plasma extracellular vesicle (EV) synaptosome-associated protein 25 (SNAP-25), growth-associated protein 43 (GAP-43) and synaptotagmin-1. P-value indicates the intra-group comparisons between baseline and the 1-year follow-up, and p for trend indicates the inter-group comparisons for changes.

|  |  | SNAP-25 | | | | GAP-43 | | | | Synaptotagmin-1 | | | |
| --- | --- | --- | --- | --- | --- | --- | --- | --- | --- | --- | --- | --- | --- |
|  |  | L, n=74 | H, n=28 | p (intra-group) | p (inter-group) | L, n=74 | H, n=28 | p (intra-group) | p (inter-group) | L, n=74 | H, n=28 | p (intra-group) | p (inter-group) |
| UPDRS-  II | Baseline | 8.26±5.80 | 9.04±4.90 | 0.530 | **<0.001** | 8.53± 5.67 | 8.32±5.33 | 0.868 | **<0.001** | 8.46±5.70 | 8.50±5.24 | 0.974 | **<0.001** |
|  | Follow-up | 10.38±6.11 | 13.36±6.72 | **0.035** |  | 10.65±6.30 | 12.64±6.53 | 0.161 |  | 10.57±6.25 | 12.86±6.58 | 0.107 |  |
| UPDRS-III | Baseline | 22.31±9.54 | 23.89±8.65 | 0.445 | 0.259 | 22.22±9.66 | 24.14±8.25 | 0.352 | 0.244 | 21.97±9.37 | 24.79±8.93 | 0.174 | 0.145 |
|  | Follow-up | 20.01±9.33 | 24.21±8.87 | **0.042** |  | 19.96±9.25 | 24.36±8.99 | **0.033** |  | 20.04±9.39 | 24.14±8.70 | **0.048** |  |
| Tremor | Baseline | 0.34±0.46 | 0.46±0.38 | 0.127 | **0.037** | 0.34±0.34 | 0.46±0.37 | 0.105 | **0.034** | 0.33±0.34 | 0.47±0.38 | 0.072 | **0.014** |
|  | Follow-up | 0.27±0.26 | 0.35±0.32 | 0.181 |  | 0.27±0.26 | 0.35±0.32 | 0.181 |  | 0.28±0.29 | 0.32±0.22 | 0.595 |  |
| AR | Baseline | 1.04±0.46 | 1.09±0.44 | 0.628 | 0.300 | 1.03±0.47 | 1.11±0.42 | 0.449 | 0.325 | 1.03±0.46 | 1.13±0.45 | 0.317 | 0.195 |
|  | Follow-up | 0.96±0.46 | 1.10±0.42 | 0.164 |  | 0.95±0.45 | 1.12±0.43 | 0.076 |  | 0.96±0.46 | 1.10±0.42 | 0.155 |  |
| PIGD | Baseline | 0.76±0.61 | 0.81±0.44 | 0.658 | **0.023** | 0.77±0.60 | 0.77±0.45 | 0.963 | **0.046** | 0.74±0.56 | 0.86±0.58 | 0.351 | **0.027** |
|  | Follow-up | 0.74±0.59 | 1.07±0.75 | **0.023** |  | 0.78±0.61 | 0.98±0.74 | 0.157 |  | 0.73±0.56 | 1.11±0.80 | **0.008** |  |
| MMSE | Baseline | 25.32±4.45 | 25.29±3.29 | 0.967 | 0.483 | 25.58±4.16 | 24.61±4.11 | 0.293 | 0.470 | 25.62±3.91 | 24.50±4.69 | 0.225 | 0.342 |
|  | Follow-up | 24.78±5.93 | 25.14±4.64 | 0.774 |  | 25.05±5.79 | 24.43±5.10 | 0.616 |  | 25.23±5.61 | 23.96±5.50 | 0.310 |  |
| MoCA | Baseline | 20.68±6.00 | 21.21±5.00 | 0.680 | 0.834 | 21.14±5.77 | 20.04±5.62 | 0.390 | 0.899 | 21.12±5.33 | 20.07±6.69 | 0.411 | 0.926 |
|  | Follow-up | 20.60±6.85 | 21.46±5.37 | 0.551 |  | 21.10±6.55 | 20.18±6.30 | 0.526 |  | 21.19±6.33 | 19.93±6.80 | 0.382 |  |

L: 2nd to 4th quartile at baseline; H: 1st quartile at baseline; UPDRS, unified Parkinson Disease rating scale; AR, akinetic rigidity; PIGD, postural instability and gait disturbance; MMSE, mini-mental status examination; MoCA, Montreal cognitive assessment.

Supplementary Figure 1. The original blot image of the representative SNAP-25, GAP-43, synaptotagmin-1, and HSP-70 in Figure 1A of the main text.

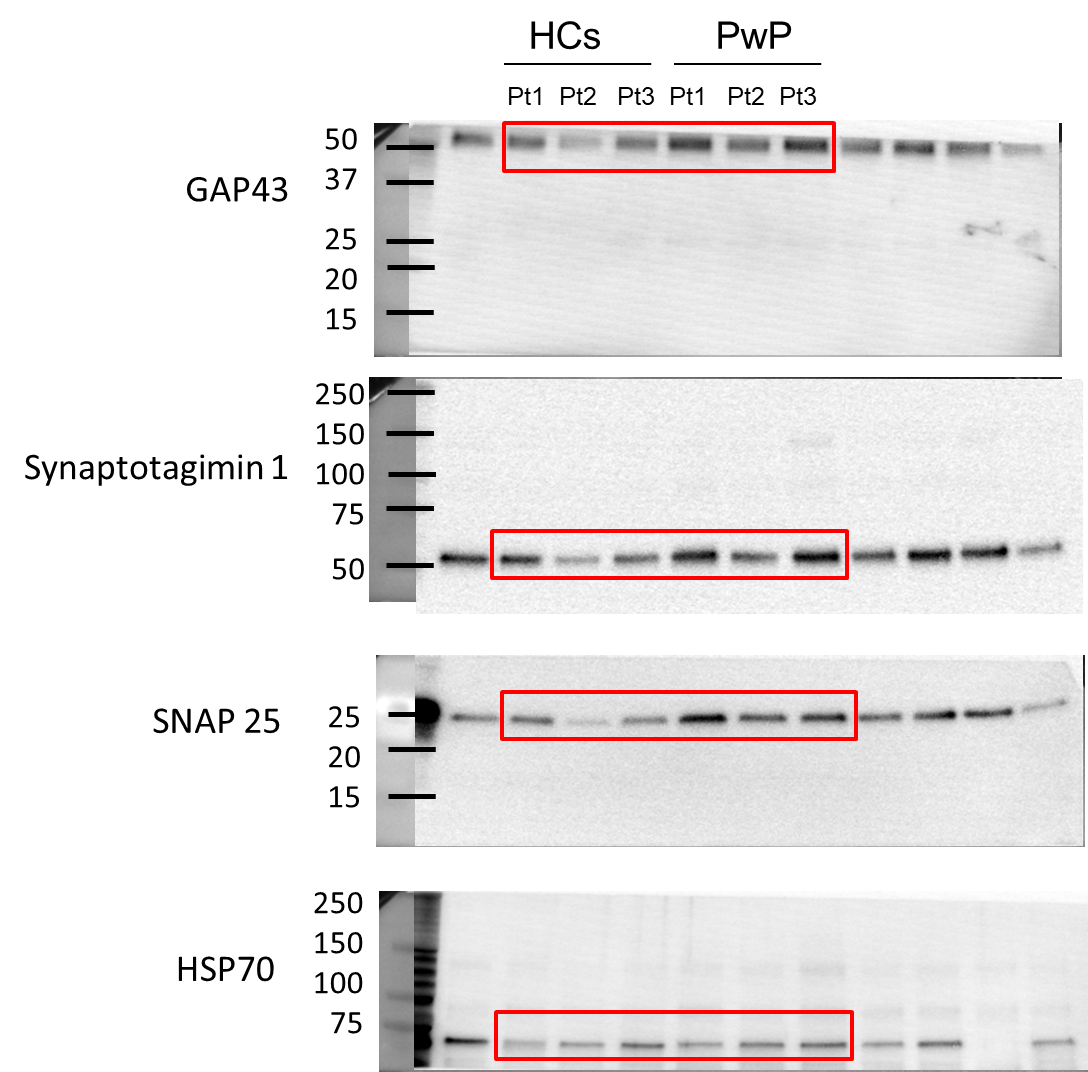
